# Digital Health Interventions and Quality of Home-based Primary Care for Older Adults: A Scoping Review Protocol

**DOI:** 10.1101/2022.04.01.22273008

**Authors:** Isis de Siqueira Silva, Cicera Renata Diniz Vieira Silva, Rayssa Horacio Lopes, Aguinaldo Jose Araujo, Renan Cabral de Figueiredo, Osvaldo de Goes Bay Junior, Luis Velez Lapao, Severina Alice da Costa Uchoa

## Abstract

**Introduction:** The use of digital health interventions has expanded, particularly in home-based primary care (HBPC) following the increase in the older adult population and the need to respond to the higher demand of chronic conditions and health frailties of this population. There was an even greater demand with COVID-19 and subsequent isolation/social distancing measures for this risk group. The objective of this study is to map and identify the uses and types of digital health interventions and their reported impacts on the quality of HBPC for older adults worldwide.

**Methods and analysis:** This is a scoping review protocol which will enable a rigorous, transparent and reliable synthesis of knowledge. The review will be developed in the theoretical perspective of Arksey and O’malley, with updates by Levac et al. and Peters et al. based on the Joanna Briggs Institute manual, and guided by the Preferred Reporting Items for Systematic Reviews and Meta-Analyses Extension for Scoping Reviews (PRISMA-ScR). Data from whiteliterature will be extracted from multidisciplinary health databases such as: the Virtual Health Library, LILACS, PubMed, Scopus, Web of Science, CINAHL and Embase; while Google Scholar will be used for gray literature. No date limit or language restrictions will be determined. The quantitative data will be analyzed through descriptive statistics and qualitative data through thematic analysis. The results will be submitted to stakeholder consultation for preliminary sharing of the study and will later be disseminated through publication in open access scientific journals, scientific events and academic and community journals. The full scoping review report will present the main impacts, challenges, opportunities and gaps found in publications related to the use of digital technologies in primary home care.

**Discussion:** The organization of this protocol will increase the methodological rigor, quality, transparency and accuracy of scoping reviews, reducing the risk of bias.

## 1 Introduction

The increase in the older adult population and the subsequent need for health systems to respond to issues of chronic diseases, weaknesses, and loss of autonomy has increased the demand for home-based primary care (HBPC) around the world. HBPC includes care that seeks to adequately meet the social and health needs of people in the residential environment. Actions are offered for promotion, prevention, minimization of disease sequelae, situations of weakness and loss of autonomy, monitoring of chronic diseases, palliative care, and support in activities of daily living. These actions can be technical, offered by health professionals or laypeople, the result of intuition, and support in daily life activities care for older adults and self-care guided by professionals (1).

The increase in life expectancy contributes to an expansion of dependence on healthcare for a long time, which includes HBPC (2). Frailty in older adults is related to a reduction of physiological factors associated with increased vulnerability, which in turn contributes to the emergence of Chronic Non-Communicable Diseases, influences the loss of functionality and is the main cause of dysfunction such as disabilities, limitation in activities or restriction in community and social participation (3,4).

An important challenge for the quality of HBPC is the need for complex coordination due to the interdependence of health services, formal and informal healthcare providers, and self-care. This coordination can be played by Primary Healthcare (PHC), hospitals or nursing services (5).

Home care is one of the PHC priorities, especially for those who cannot easily commute to health services (6). Studies about PHC and HBPC articulation present advantages such as providing users with mechanisms to access longitudinal care and promoting improved quality of care with lower costs due to a stronger relationship between the person and their caregiver (7,8). Expanding coverage and quality of services are of paramount importance for PHC (9) as a strategy to reorganize health systems in order to guarantee longitudinal and comprehensive care for chronic patients in the territories covered, especially in cases where HBPC is the timeliest form of care (5,9).

HBPC demand has increased significantly during the COVID-19 pandemic, considering that older adults, carriers of chronic diseases and affected by immunosenescence, are more susceptible to infectious disease (7). HBPC was used to reduce attendance at emergency services and ensure that chronic medical problems were treated within the home environment to prevent their worsening (10,11), and digital health was used in this same perspective.

Faced with the challenges in PHC from COVID-19, the use of Information and Communication Technologies gained even more prominence due to the operability and versatility of generating information at an opportune time (12,13,14). Digital health assists healthcare workers in diagnosing, monitoring, and communicating with older adult patients around the world, especially during the COVID-19 pandemic (15). Digital health can be defined as a safe and cost-effective way of using information and communication technologies in health and related areas (16). It is an umbrella term which includes a series of services and systems, such as medical and applied health informatics, teleconsultations, telemonitoring, e-learning and mHealth. Its use can contribute to strengthen health systems by quickly making reliable and up-to-date health information available (17).

The use of digital health at home improves access to healthcare, enhances a sense of security and often reduces commuting (18,19). A study conducted in Indonesia identified needs and opportunities for enabling the use of cell phones and mobile applications for the health of older adults (15). There is also an increase in the use of digital tools for people with chronic conditions (20), even in PHC (13). The expanded use of digital health indicates that there is a field of research to be explored. Therefore, the relevance of carrying out studies that emphasize HBPC for older adults mediated by digital health in the context of PHC is evidenced.

Given the importance of the relationship between digital health and HBPC in different countries, especially strengthened in the current pandemic period and its possibilities for its continuity even after the pandemic due to the significant benefits, it is fundamental to retrieve and systematize the evidence of these experiences and it is impacts on the quality of healthcare. This is particularly relevant for groups of older adults among whom chronic conditions are prevalent and who are susceptible to HBPC.

This study uses a Donabedian model as a theoretical framework quality assessment in health (21,22). This concept of quality applied to healthcare is, in practice, approached using a set of desirable attributes, which are called (the seven) pillars of quality: efficacy, effectiveness, efficiency, optimization, acceptability, legitimacy and equity (21,22).

A preliminary search was conducted in January 2022 in the PROSPERO, MEDLINE, Cochrane Database of Systematic Reviews, and Joanna Briggs Institute (JBI) Evidence Synthesis databases; no review or research protocol with a similar theme applied to PHC was identified.

Thus, the objective of this study is to map and identify the uses and types of health interventions and their impacts on the digital quality of HBPC for older adults worldwide.

## 2 Materials and Methods

This study is a scoping review protocol which seeks to answer broader research questions. The study will identify and map emerging evidence on the topic addressed, synthesizing knowledge with rigor, transparency and reliability. It is based on JBI criteria guided by the theoretical framework of Arksey and O’malley (23), with updates from Levac et al. (24) and Peters et al. (25), as well as by the Preferred Reporting Items for Systematic Reviews and Meta-Analyses Extension for Scoping Reviews (PRISMA-ScR) (26). The protocol was registered in the Open Science Framework (OSF) (https://osf.io/vgkhy). This review will be conducted in six steps, as shown in Figure 1: formulating the research question, identifying relevant studies, selecting studies, extracting and coding data, analyzing and interpreting results, and consulting stakeholders (27).

**Figure 1:**
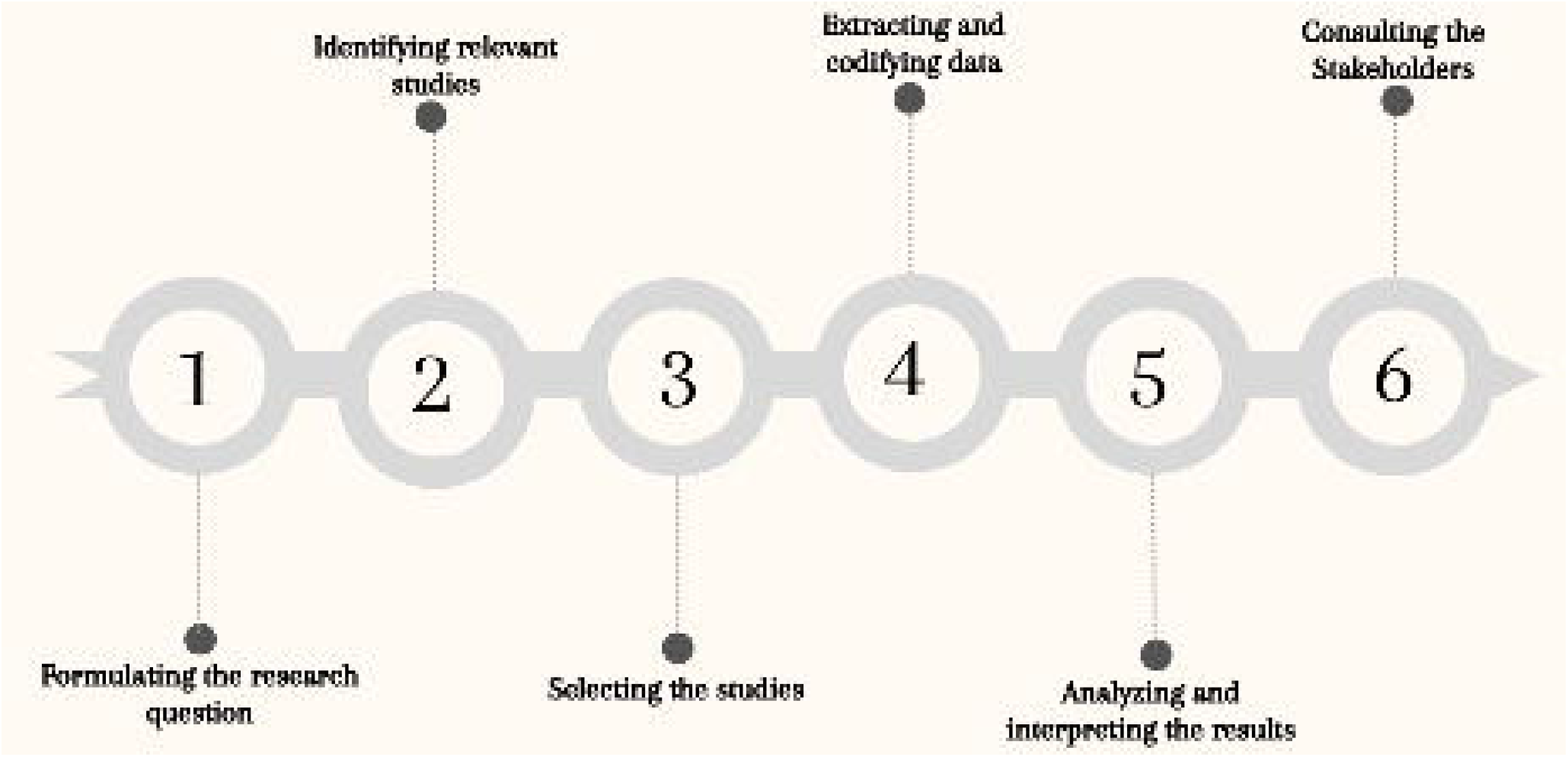
Steps of the Scoping Review (27): **Source**: prepared by the authors, 2022.

### 2.1 Step 1: Formulating the research question

The research questions were formulated through the PCC mnemonic conceptual model – (Population, Concept, Context) (26), as:

**P**: Older adults;

**C**: Digital health interventions;

**C**: Home-based primary care.

The following research questions were prepared by the authors according to the PCC:

1. Which countries use digital health interventions in home-based primary care for older adults?
2. What sort of digital health interventions are used in home-based primary care for older adults?
3. What is the measured impact of digital health interventions on the quality of home-based primary care for older adults?

The key concepts for elaborating the research questions are described in Table 1.

**Table 1.** Key concepts for the study questions.

| Concept | Definition |
| --- | --- |
| Older adult | For the World Health Organization (28), the concept of “old age” is multidimensional, and includes the terms chronological (based on the date of birth), biological (related to the capacity of the human body), psychological (related to the psycho-emotional functioning) and social age (related to social roles). For the United Nations (29), the definition of an older adult is related to those who are 60 years old or more, but at the same time they affirm that there is a diversity of older people with different needs, abilities, lifestyles, experiences and preferences which are influenced by age, gender, health, income, education, ethnicity and other factors. |
| Digital health interventions | The classification of digital health interventions (DHIs) categorizes the different ways in which digital technologies are being used to support health system needs. This classification framework is primarily targeted at public health audiences, and aims to promote an accessible and bridging language for health program planners to articulate functionalities of |
|  | digital health implementations (19). |
| HBPC (home-based primary care) | This is a model of providing long-term primary care at home, ranging from palliative care, rehabilitation, and disease management to care coordination. The multidisciplinary team has older adults with chronic diseases and physical and cognitive disabilities as its main clientele (30,31). |
| Care quality | The quality of care can be defined in three dimensions: technical (accuracy in the choice of actions and the way in which they are produced), interpersonal (social and psychological relationships between care providers and users) and organizational (conditions in which services are offered comprehensively and with continuity of care, coverage, coordination of actions, access and accessibility to services (21). |

**Source:** prepared by the authors, 2022.

### 2.2 Step 2: Identifying relevant studies

The following steps will be taken to enhance the identification of documents in white literature and gray literature:

#### 2.2.1 Exploratory Search

The initial search was conducted in PubMed using Medical Subject Headings (MeSH) in English to identify main descriptors, synonyms, and keywords included in titles, abstracts, and indexed terms of publications regarding the theme. A similar search was conducted in Portuguese using the Virtual Health Library (VHL) and *Descritores em Ciências da Saúde* (*DeCS*).

Moreover, a librarian improved the search strategy using four controlled vocabularies (*DeCS*, MESH, EMTREE, and THESAURUS CINAHL’s) to obtain a wide range of multidisciplinary results in different databases. Natural language (non-controlled vocabulary) was also used to increase the sensitivity of the strategy (32).

The search strategy was constructed using the Extraction, Conversion, Combination, Construction, and Use model, which enables developing highly sensitive search strategies by following a set of complementary steps (32).

#### 2.2.2 Search strategy

English was used to structure the research strategy, considering that it is the main language used in the scientific environment (33). Chart 1 organizes the main descriptors available in the DeCS that started the search strategy carried out by the authors based on the PCC, the standard search strategy is available in Appendix I. The detailed search strategy for all data sources (i.e. white and gray literature) will be attached to the final scoping review.

**Chart 1**: Descriptors used according to the PCC Mnemonic.

| Mnemonic | Descriptor | Synonyms/keywords |
| --- | --- | --- |
| **P** | **aged / elderly / frail elderly** | Older adults; Aging; older persons; elderly care; aged people; Older Adult |
| **C** | **Digital health** | eHealth; e-Health; telehealth; Telecare; mHealth; Telerehabilitation; Telehomecare; home telehealth; Home telecare; telemonitoring; telecare monitoring system; telenursing; Digital Health; Digital Health Strategies; Digital Health Strategy; Digital Health Interventions; eHealth Strategies and Policies; Telemedicine |
| **C** | **Home-based care services** | home health services; home monitoring; home health care; home care; home-based primary care; Hospitalization at home; Home-based care; Home healthcare; Home-based primary care |

**Source:** prepared by the authors, 2022.

#### 2.2.3 Data sources

The data collection will be conducted in the following indicated portals and databases: LILACS; PubMed; Scopus; Web of Science; CINAHL and Embase. Gray literature will be searched through Google Scholar and preprint repositories. The appropriate strategy will be applied to each of them, and the title and abstract of all identified studies will be evaluated and the duplicates removed.

The search strategy was pre-tested on PubMed for white literature (Appendix II) and Google Scholar for gray literature (Appendix III) to check for the possibility of data collection limitations related to the search strategy.

#### 2.2.4 Additional Sources

Reference lists of included studies will be consulted for verification of additional publications. If needed, corresponding authors will be contacted via e-mail for additional information.

### 2.3 Step 3: Selecting the studies

The study selection process will be guided by the steps proposed in the Preferred Reporting Items for Systematic Review and Meta-Analyses (PRISMA-P) for both white and gray literature, which are: (1) identification; (2) screening; (3) eligibility; and (4) inclusion, which will be presented in detail in the review selection diagram.

The selection process of publications belonging to the gray literature will follow the guidelines recommended by Godin et al. (34), with specific strategies for searches on Google Scholar and Preprints repositories. Combinations of the following groups of search terms will be used: Aged OR elderly OR “middle age” OR “old people” OR “very elderly” AND Digital Health OR Telemedicine OR teleconsultation OR “electronic consultation” OR “remote consultation” OR telehealth “home health care” OR “home care”. The search terms and the number of results retrieved for each gray literature search strategy will be recorded and will follow the other proposed selection steps. The results from Google Scholar will be sorted by relevance and the first hundred will be included in the screening (34).

Identified studies will be grouped in the Endnote reference manager and duplicates removed. The Rayyan software program will be used in the evaluation of studies by titles and abstracts to assist in blinding the reviewers (35) and any differences between the two reviewers (IdSS and AJA) will be discussed with a third reviewer (SACU). Studies selected by title and abstract will be retrieved in full and exported to a database in the Microsoft Excel^®^ program. After reading the full text and building the final review sample, data will be extracted by the two independent reviewers, highlighting all reasons for exclusion when necessary and the entire selection process, eligibility, inclusion and reasons for exclusions will be presented in a specific flowchart (24).

A pilot test will be carried out with all authors of the protocol before starting data collection in order to ensure alignment in the selection process. Each author will randomly sample 25 titles and abstracts in one data source, then screen them using eligibility criteria. Afterwards, the team will meet to discuss the discrepancies and make necessary changes to the criteria and definitions. Screening will only start when 75% or more similarity is achieved (27).

#### 2.3.1 Inclusion criteria

Publications that address the use of digital health interventions in HBPC for older adults will be included, available in full, which answer the study questions.

The following will be included:

a. Primary studies.
b. Gray literature, including government manuals, as well as dissertations and theses.

Time filters will not be applied to the searches, as the search strategies will contain descriptors and terms referring to digital health. The search will not be limited by date or language.

#### 2.3.2 Exclusion criteria

Duplicate publications, literature reviews, theoretical essays, editorials, expert opinions and brief communications will be excluded.

### 2.4 Step 4: Extracting and codifying data

Data will be extracted according to appendix IV and included if they align with the objectives and research questions of the scoping review. Data related to the included studies will be extracted by two independent reviewers to reduce the chance of errors and biases using a data extraction form based on the JBI model and adapted by the authors. The instrument can receive updates during the research to obtain a deeper understanding of the theme, as according to Peters et al. (25).

### 2.5 Step 5: Analyzing and interpreting the results

Descriptive statistics (absolute and percentage frequencies) will be used to analyze quantitative data with the help of the Microsoft Excel^®^ program. Qualitative data analysis will be guided by thematic analysis (36).

This step will be divided into three others, according to Levac (24), namely: (1) data analysis; (2) exposure of results linked to research questions; and (3) interpreting the implications of the results for other research and services.

A map of identified countries that use digital health interventions in HBPC for older adults will be developed using the GeoDa version 1.20 software program (Center for Spatial Data Science, Chicago, IL, USA).

All results will be discussed with the relevant literature. The evidence synthesis will be presented in a descriptive format through tables, diagrams, and thematic maps to better visualize the results found. A narrative summary will follow the mapped data, and report how the results relate to the review objective and questions.

#### 2.5.1 Summary of evidence, conclusions, implications of findings

The main results will be summarized (including an overview of the concepts, themes and types of evidence available), the research questions and the objective should be answered based on the results found. Expectations about the implications of the findings on digital health interventions and their relevance to the home-based care of older adults will be presented.

### 2.6 Step 6: Consulting the Stakeholders

The final report guided by the PRISMA-ScR (26) will include the results in flowcharts, charts, or figures, and will be presented to a group of five (5) stakeholders with experience in digital health aimed at home-based care belonging to the faculty of the university to which the authors of this study are linked, in the Postgraduate Program in Public Health (PPGSCol).

The procedure will include sending an individual invitation to candidates for research participants, explaining the purpose of their participation and, if they accept, they will sign the Free and Informed Consent Form. Preliminary results and informed consent will be included in an electronic form and sent to stakeholders via e-mail. Stakeholders will not be identified, and authors will request the appreciation of results and possible new fields or evidence.

The objectives of this strategy will be the preliminary sharing of study findings, being considered a mechanism for knowledge transfer and exchange, as well as to develop effective dissemination strategies and ideas for future studies (24).

## 3 Ethics and dissemination of the results

The study does not directly involve patients, but the stakeholder consultation was approved by the Research Ethics Committee of the Onofre Lopes University Hospital/Federal University of Rio Grande do Norte CAEE 54853921.0.0000.5292. The results will be presented at scientific conferences, events with stakeholders and submitted for open-access publication in a peer-reviewed journal.

## 4 Discussion

This protocol was developed by researchers trained in this type of research and following the methodological criteria suggested by Arksey and O’Malley (23), Levac et al. (24), and JBI (27). The organization of this protocol will increase the methodological rigor, quality, transparency and accuracy of scoping reviews, reducing the risk of bias. Scoping review protocols contribute to an increasing need to synthesize and summarize research following a reproducible design, implementation and reporting method (37).

The scoping review will be able to present the convergence of two emerging themes, namely, digital health, which offers an opportunity to address health system challenges, improve coverage and maintain the quality of service (38) and home primary care for older adults who demand continuous and sustainable long-term care (7).

## 5 Strengths and limitations

The methodological rigor adopted in this protocol, as well as the training and experience of the researchers, will ensure quality and transparency for the development of the scoping review. In addition, this is the first study to propose mapping the use and type of digital health interventions used, and their impacts on the quality of PHC-coordinated home care for older adults.

However, two limitations in the search strategy can be highlighted. The first is that the definition of Digital Health is recent (2020) and evolving. If there are changes in WHO definitions of digital health by the study selection stage, the terms will be updated. The second is the structuring of the search strategy in English which may not include publications from the gray literature in the native language of some countries. Thus, the search strategy may be adapted to Portuguese, Spanish, and French to extend the reach. Therefore, we constructed the search in a manner that increased comprehensiveness to minimize the effect of these limitations.

## 6 Conclusion

The present protocol is proposed to guide a scoping review that will map and identify the uses and types of health interventions and their impacts on the digital quality of HBPC for older adults worldwide. The proposed study will provide a reliable source of evidence for managers, digital tool developers, healthcare professionals, caregivers, patients, and future research to guide the use of digital health interventions in the practice of HBPC for older people, and its results may guide discussions for the elaboration of upcoming healthcare policies and guidelines.

## Data Availability

All data produced in the present work are contained in the manuscript

https://osf.io/vgkhy/

http://creativecommons.org/licenses/by-nc/4.0/

## 7 Footnotes

### 7.1 Contributors

SACU proposed the study and coordinated the elaboration of the protocol. IdSS developed the protocol. IdSS, CRDVS, RHL, OdGBJ, AJA, RCF, LVL and SACU participated in the discussion of the theoretical and methodological aspects of the study. CRDVS, IdSS and RHL conducted the pilot searches to substantiate the search strategy. All authors reviewed the protocol and approved its final version for publication.

### 7.4 Patient consent for publication

Not required

## 7.5 Acknowledgments

The preprint of this work was posted on medRxiv (39).

## APPENDIX I Standard review search strategy

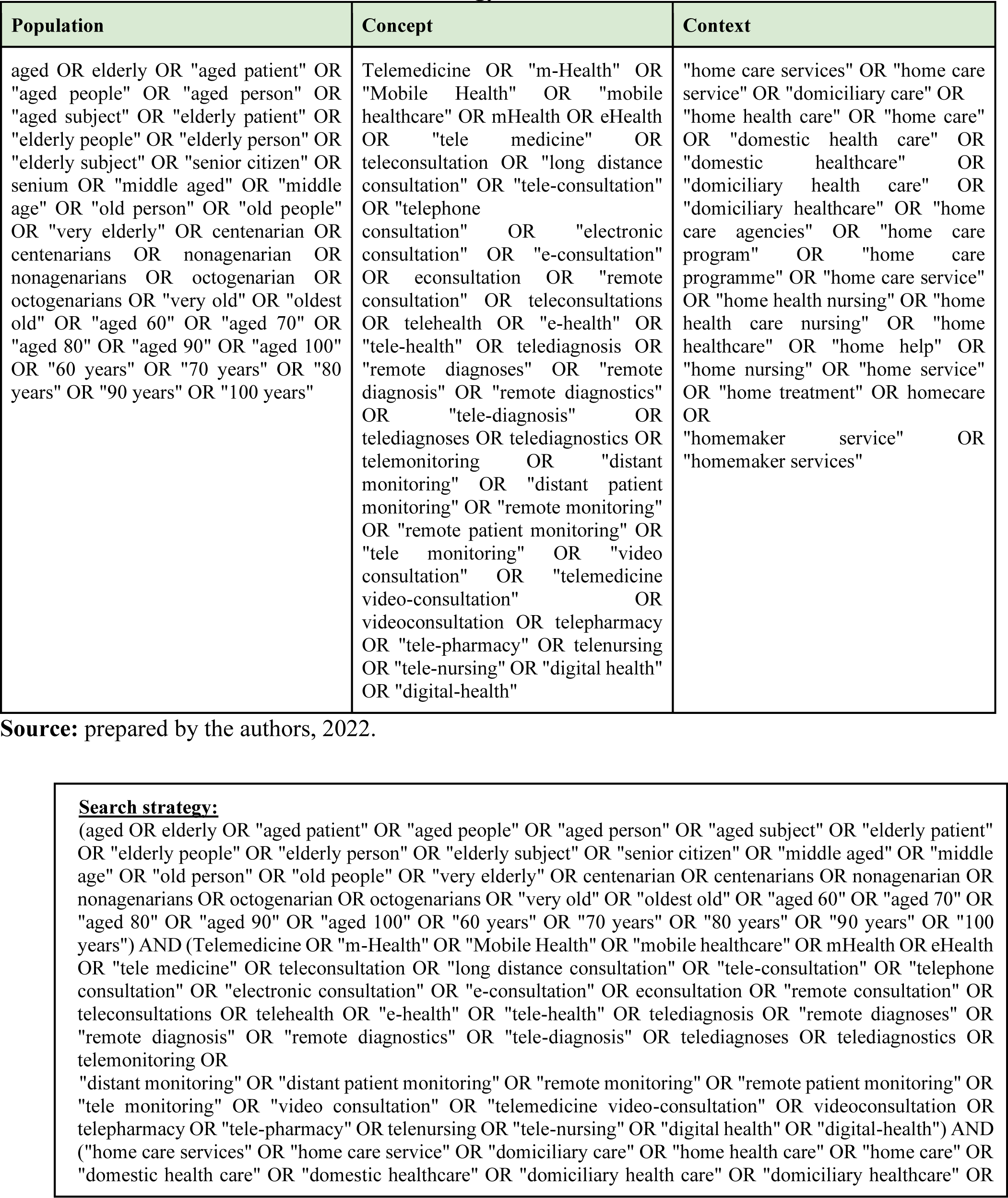
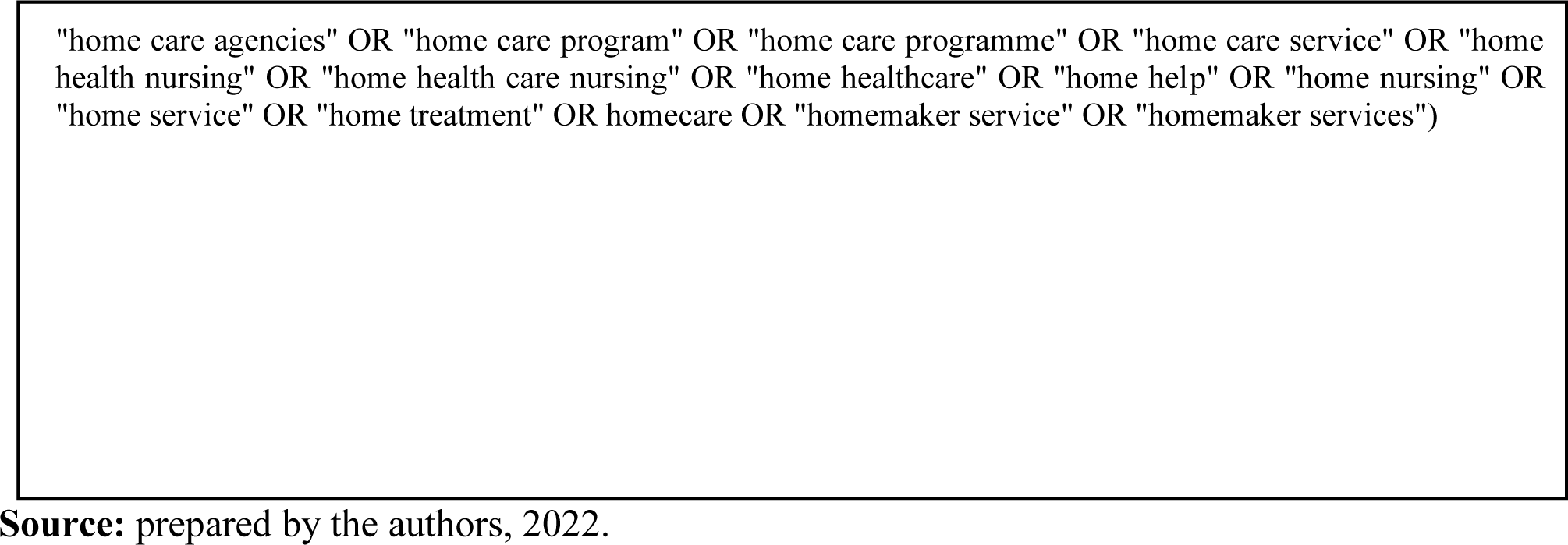

## APPENDIX II Search Strategy PubMed

Search performed in the PubMed database on July 21, 2022.

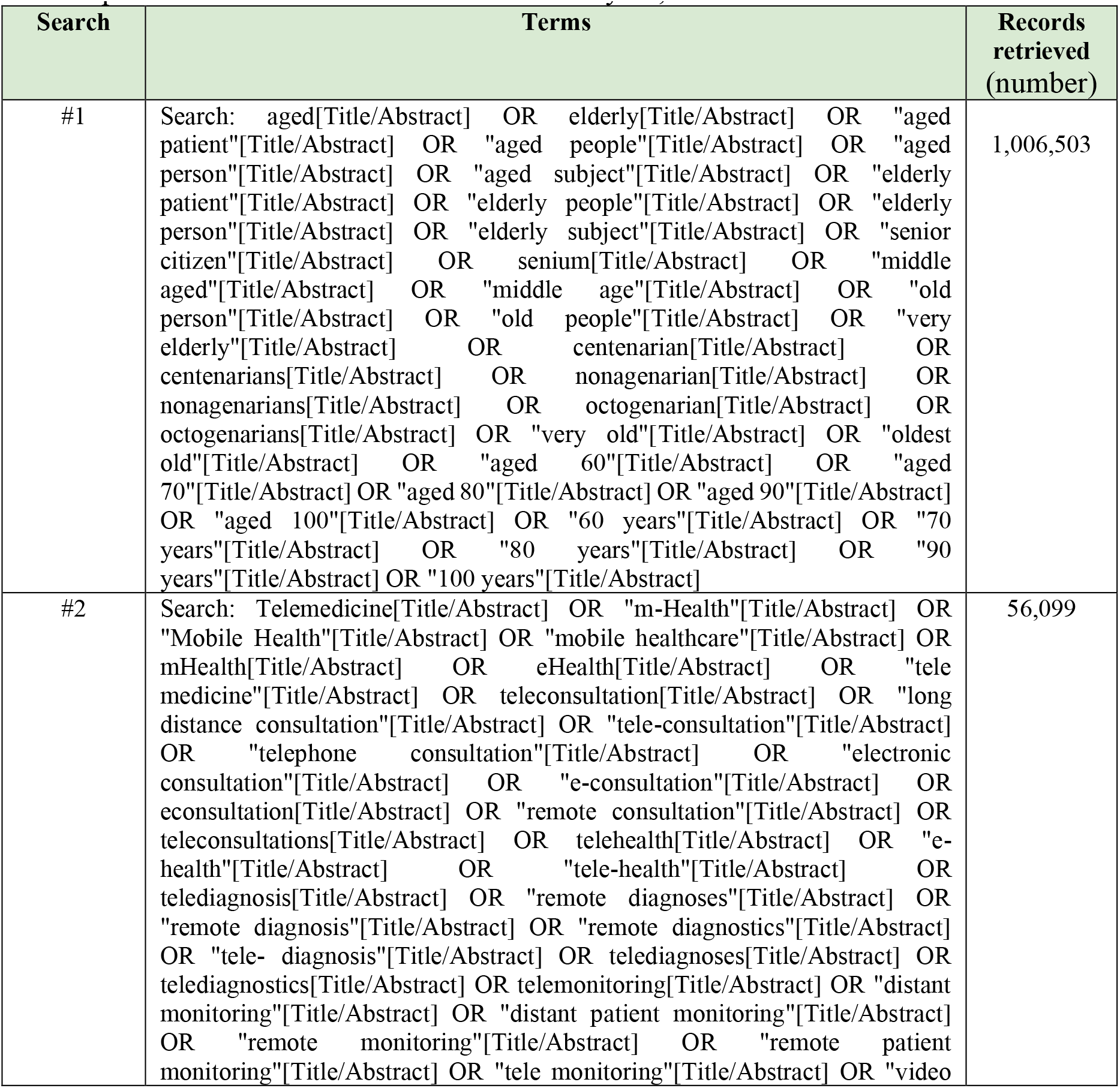
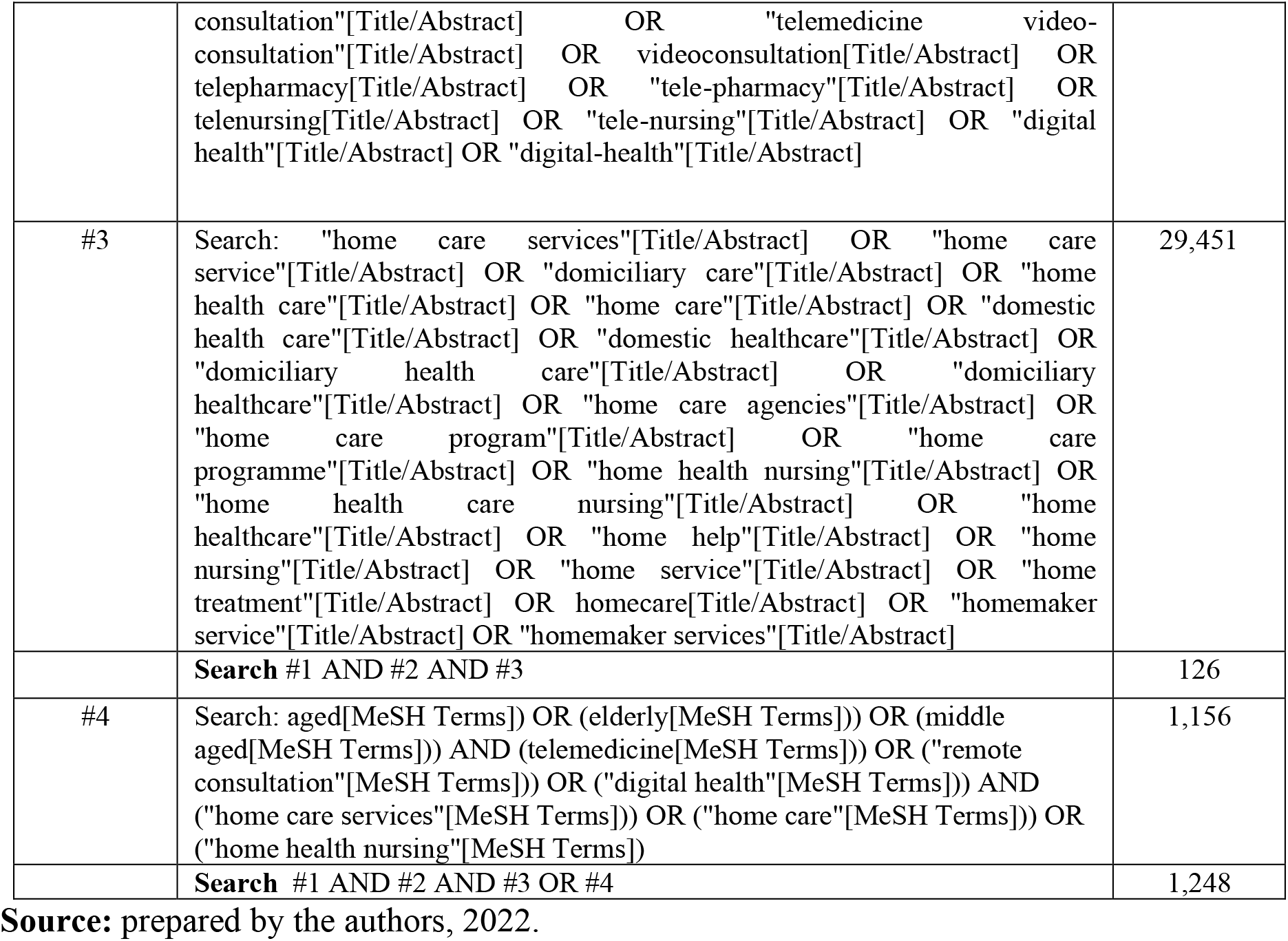

| Search | Terms | Records retrieved (number) |
| --- | --- | --- |
| #1 | Search: aged[Title/Abstract] OR elderly[Title/Abstract] OR "aged patient"[Title/Abstract] OR "aged people"[Title/Abstract] OR "aged person"[Title/Abstract] OR "aged subject"[Title/Abstract] OR "elderly patient"[Title/Abstract] OR "elderly people"[Title/Abstract] OR "elderly person"[Title/Abstract] OR "elderly subject"[Title/Abstract] OR "senior citizen"[Title/Abstract] OR senium[Title/Abstract] OR "middle aged"[Title/Abstract] OR "middle age"[Title/Abstract] OR "old person"[Title/Abstract] OR "old people"[Title/Abstract] OR "very elderly"[Title/Abstract] OR centenarian[Title/Abstract] OR centenarians[Title/Abstract] OR nonagenarian[Title/Abstract] OR nonagenarians[Title/Abstract] OR octogenarian[Title/Abstract] OR octogenarians[Title/Abstract] OR "very old"[Title/Abstract] OR "oldest old"[Title/Abstract] OR "aged 60"[Title/Abstract] OR "aged 70"[Title/Abstract] OR "aged 80"[Title/Abstract] OR "aged 90"[Title/Abstract] OR "aged 100"[Title/Abstract] OR "60 years"[Title/Abstract] OR "70 years"[Title/Abstract] OR "80 years"[Title/Abstract] OR "90 years"[Title/Abstract] OR "100 years"[Title/Abstract] | 1,006,503 |
| #2 | Search: Telemedicine[Title/Abstract] OR "m-Health"[Title/Abstract] OR "Mobile Health"[Title/Abstract] OR "mobile healthcare"[Title/Abstract] OR mHealth[Title/Abstract] OR eHealth[Title/Abstract] OR "tele medicine"[Title/Abstract] OR teleconsultation[Title/Abstract] OR "long distance consultation"[Title/Abstract] OR "tele-consultation"[Title/Abstract] OR "telephone consultation"[Title/Abstract] OR "electronic consultation"[Title/Abstract] OR "e-consultation"[Title/Abstract] OR econsultation[Title/Abstract] OR "remote consultation"[Title/Abstract] OR teleconsultations[Title/Abstract] OR telehealth[Title/Abstract] OR "e-health"[Title/Abstract] OR "tele-health"[Title/Abstract] OR telediagnosis[Title/Abstract] OR "remote diagnoses"[Title/Abstract] OR "remote diagnosis"[Title/Abstract] OR "remote diagnostics"[Title/Abstract] OR "tele- diagnosis"[Title/Abstract] OR telediagnoses[Title/Abstract] OR telediagnosics[Title/Abstract] OR telemonitoring[Title/Abstract] OR "distant monitoring"[Title/Abstract] OR "distant patient monitoring"[Title/Abstract] OR "remote monitoring"[Title/Abstract] OR "remote patient monitoring"[Title/Abstract] OR "tele monitoring"[Title/Abstract] OR "video | 56,099 |
|  | consultation"[Title/Abstract] OR "telemedicine video-consultation"[Title/Abstract] OR videoconsultation[Title/Abstract] OR telepharmacy[Title/Abstract] OR "tele-pharmacy"[Title/Abstract] OR telenursing[Title/Abstract] OR "tele-nursing"[Title/Abstract] OR "digital health"[Title/Abstract] OR "digital-health"[Title/Abstract] |  |
| #3 | Search: "home care services"[Title/Abstract] OR "home care service"[Title/Abstract] OR "domiciliary care"[Title/Abstract] OR "home health care"[Title/Abstract] OR "home care"[Title/Abstract] OR "domestic health care"[Title/Abstract] OR "domestic healthcare"[Title/Abstract] OR "domiciliary health care"[Title/Abstract] OR "domiciliary healthcare"[Title/Abstract] OR "home care agencies"[Title/Abstract] OR "home care program"[Title/Abstract] OR "home care programme"[Title/Abstract] OR "home health nursing"[Title/Abstract] OR "home health care nursing"[Title/Abstract] OR "home healthcare"[Title/Abstract] OR "home help"[Title/Abstract] OR "home nursing"[Title/Abstract] OR "home service"[Title/Abstract] OR "home treatment"[Title/Abstract] OR homecare[Title/Abstract] OR "homemaker service"[Title/Abstract] OR "homemaker services"[Title/Abstract] | 29,451 |
|  | **Search #1 AND #2 AND #3** | 126 |
| #4 | Search: aged[MeSH Terms]) OR (elderly[MeSH Terms])) OR (middle aged[MeSH Terms])) AND (telemedicine[MeSH Terms])) OR ("remote consultation"[MeSH Terms])) OR ("digital health"[MeSH Terms])) AND ("home care services"[MeSH Terms])) OR ("home care"[MeSH Terms])) OR ("home health nursing"[MeSH Terms])) | 1,156 |
|  | **Search #1 AND #2 AND #3 OR #4** | 1,248 |

**Source:** prepared by the authors, 2022.

## APPENDIX III Search Strategy for gray literature in the Google Scholar

*Search performed in Google Scholar on July 20, 2022.

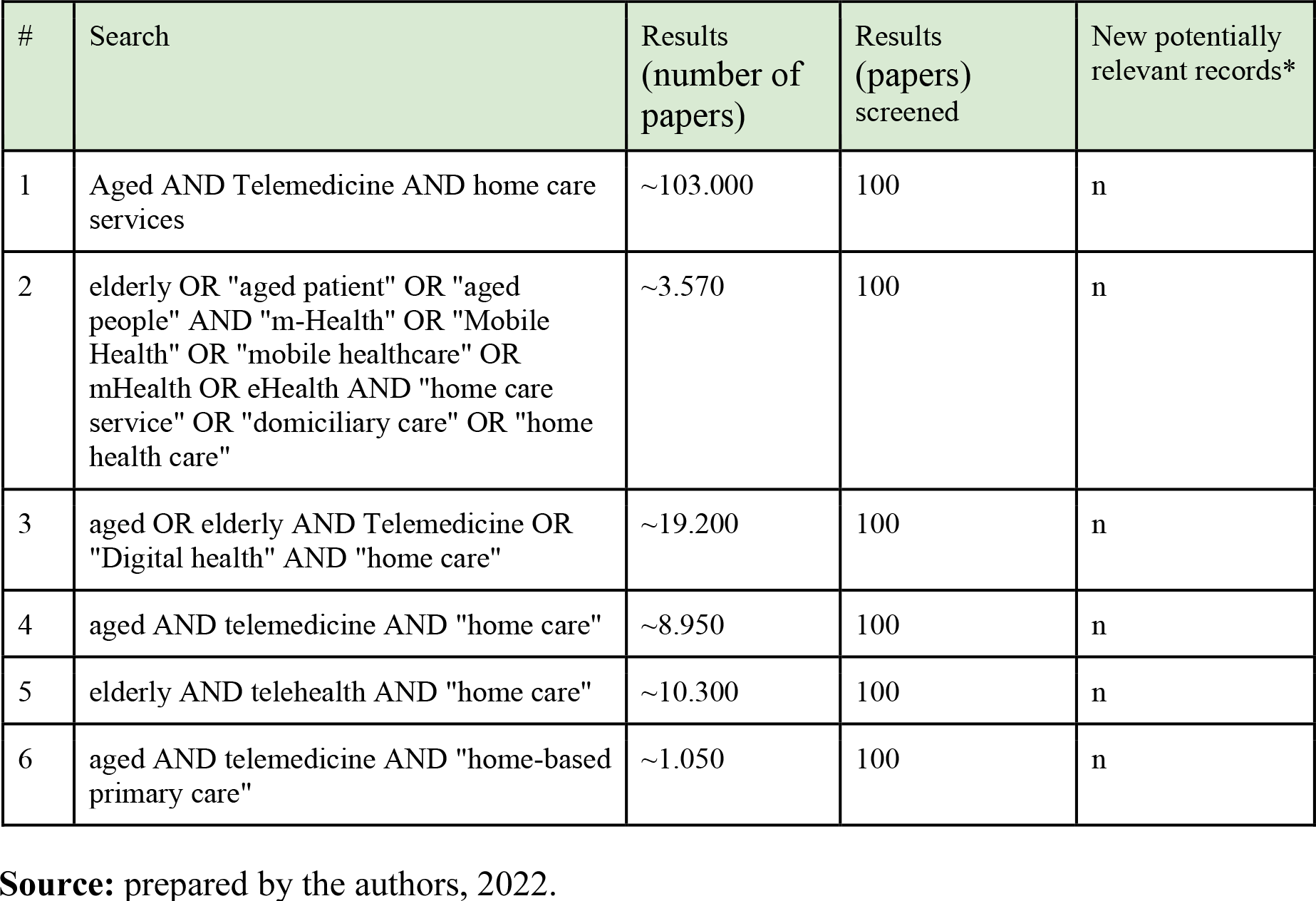

## APPENDIX IV Standard Data Collection Instrument

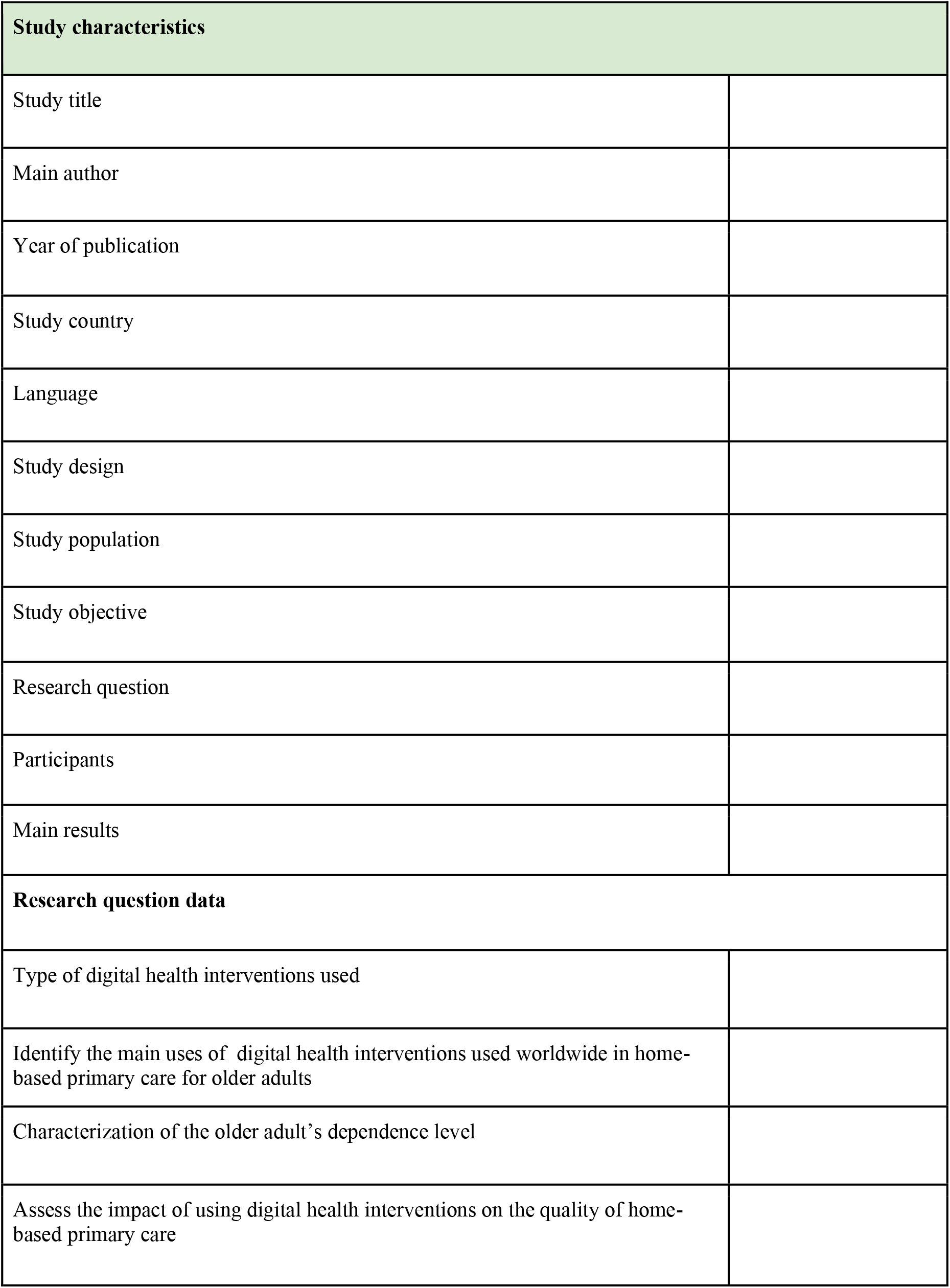
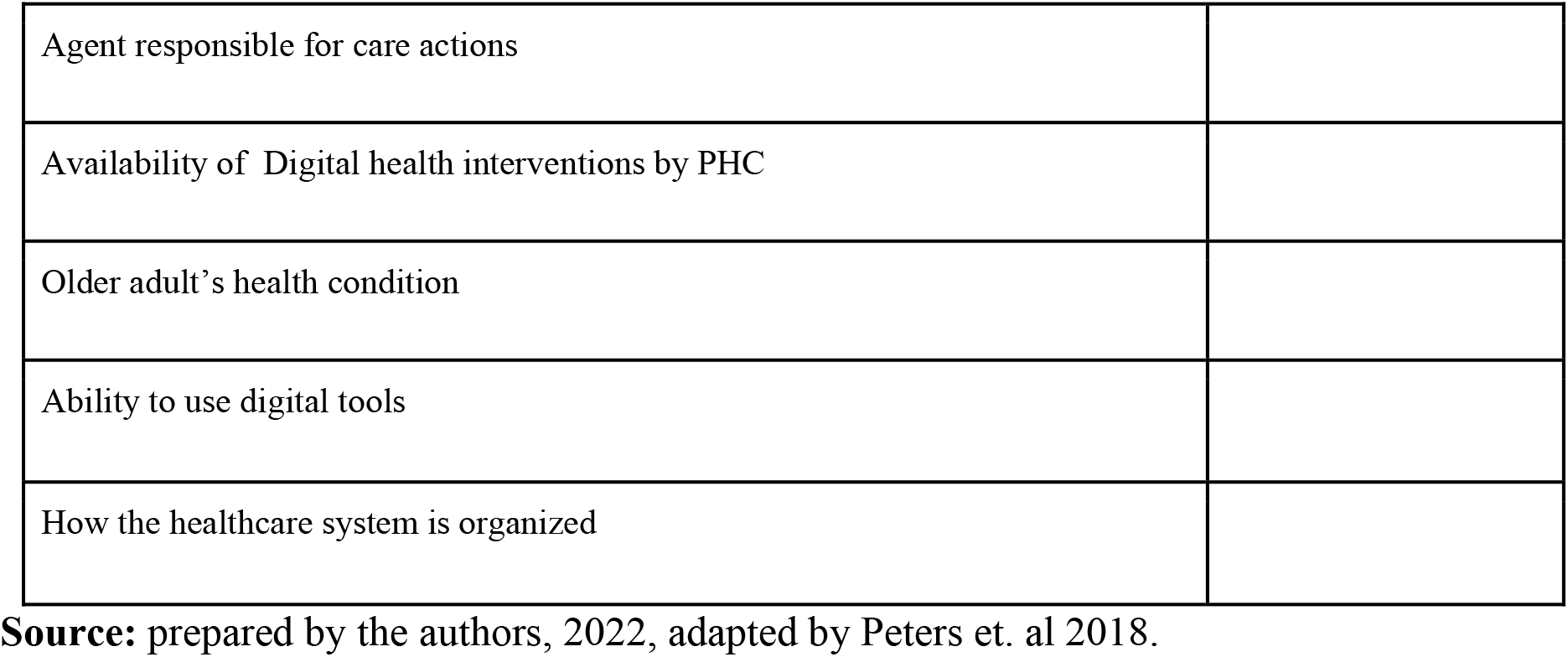

APPENDIX IV – Standard Data Collection Instrument
|  |
| --- |
| **Study characteristics** |
| Study title |
| Main author |
| Year of publication |
| Study country |
| Language |
| Study design |
| Study population |
| Study objective |
| Research question |
| Participants |
| Main results |
| **Research question data** |
| Type of digital health interventions used |
| Identify the main uses of digital health interventions used worldwide in home-based primary care for older adults |
| Characterization of the older adult's dependence level |
| Assess the impact of using digital health interventions on the quality of home-based primary care |
| **Running Title** |
| Agent responsible for care actions |
| Availability of Digital health interventions by PHC |
| Older adult’s health condition |
| Ability to use digital tools |
| How the healthcare system is organized |

**Source:** prepared by the authors, 2022, adapted by Peters et. al 2018.

