## Supplementary material for "Digital Health Interventions and Quality of Home-based Primary Care for Older Adults: A Scoping Review Protocol": Checklist prisma: S2_Checklist_PRISMA_ScR.docx (from another version of this preprint: version 1, 2022-04-05)

**S1 Checklist. Preferred Reporting Items for Systematic reviews and Meta-Analyses extension for Scoping Reviews (PRISMA-ScR) Checklist**

| **SECTION** | **ITEM** | **PRISMA-ScR CHECKLIST ITEM** | **REPORTED ON PAGE #** |
| --- | --- | --- | --- |
| **TITLE** | | | |
| Title | 1 | Digital health and quality of home-based primary care for older adults: A scoping review protocol | 1 |
| **ABSTRACT** | | | |
| Structured summary | 2 | Abstract provided | 2 |
| **INTRODUCTION** | | | |
| Rationale | 3 | Given the importance of the digital health and HBPC relationship, especially enhanced in the current pandemic period and its possibilities for its continuity even after the pandemic due to the significant benefits, it is of fundamental importance to retrieve and systematize the evidence of these experiences. | 4 |
| Objectives | 4 | This study propose a scoping review protocol to map the uses and types of digital health strategies and analyze their impacts on the quality of HBPC for older adults around the world. | 4 |
| **METHODS** | | | |
| Protocol and registration | 5 | A protocol was elaborated and registered on the Open Science Framework and can be accessed using the following link <<https://osf.io/vgkhy/>> | 5 |
| Eligibility criteria | 6 | Only publications which address digital health strategies, used worldwide in the context of home-based care for older adults, available in full, which answer the study questions (Table 2), and address information on how digital health strategies are being used in home-based primary care for older adults will be considered. | 11 |
| Information sources* | 7 | The search for evidence was performed in the following indicated portals and databases: LILACS; PubMed; Scopus; Web of Science; CINAHL and Embase. Gray literature will be searched through Google Scholar and preprint repositories. | 10 |
| Search | 8 | Search strategy provided | Appendix 1 |
| Selection of sources of evidence† | 9 | Two researchers will independently evaluate the articles for inclusion, examining the titles, abstracts and full texts of the studies returned during the search process, using Rayyan software | 10 |
| Data charting process‡ | 10 | Data will be extracted using a worksheet constructed by authors. | Appendix 2 |
| Data items | 11 | - Study title - Main author - Year of publication - Study country - Language - Study design - Study population - Study objective - Research question - Participants - Main results - Research question data - Type of digital health strategy used - Identify the main uses of digital health strategies used worldwide in home-based primary care for older adults - Characterization of the older adult’s dependence level - Assess the impact of using digital health strategies on the quality of home-based primary care - Agent responsible for care actions - Availability of digital health strategies by PHC - Older adult’s health condition - Ability to use digital tools - How the healthcare system is organized | Appendix 2 |
| Critical appraisal of individual sources of evidence§ | 12 | No applicable | - |
| Synthesis of results | 13 | This review will generate quantitative and qualitative data that will be analyzed according to proper techniques for each type of data. Quantitative data will be synthesized and analyzed using simple descriptive statistics. Alternatively, qualitative data will be analyzed using thematic analysis. | 12 |
| **RESULTS** | | | |
| Selection of sources of evidence | 14 | not applicable | N/A |
| Characteristics of sources of evidence | 15 | not applicable | N/A |
| Critical appraisal within sources of evidence | 16 | not applicable | N/A |
| Results of individual sources of evidence | 17 | not applicable | N/A |
| Synthesis of results | 18 | not applicable | N/A |
| **DISCUSSION** | | | |
| Summary of evidence | 19 | Summarized results will be presented to five stakeholders with experience in digital health strategies aimed at home-based care. The objectives of this strategy will be the preliminary sharing of study findings, being considered a mechanism for knowledge transfer and exchange, as well as to develop effective dissemination strategies and ideas for future studies. | 13 |
| Limitations | 20 | A large number of publications in the gray literature may imply an important limitation of the study but also meaning that digital home care still lacks maturity. | 3 |
| Conclusions | 21 | not applicable | N/A |
| **FUNDING** | | | |
| Funding | 22 | no funding | - |

JBI = Joanna Briggs Institute; PRISMA-ScR = Preferred Reporting Items for Systematic reviews and Meta-Analyses extension for Scoping Reviews.

* Where *sources of evidence* (see second footnote) are compiled from, such as bibliographic databases, social media platforms, and Web sites.

† A more inclusive/heterogeneous term used to account for the different types of evidence or data sources (e.g., quantitative and/or qualitative research, expert opinion, and policy documents) that may be eligible in a scoping review as opposed to only studies. This is not to be confused with *information sources* (see first footnote).

‡ The frameworks by Arksey and O’Malley (6) and Levac and colleagues (7) and the JBI guidance (4, 5) refer to the process of data extraction in a scoping review as data charting*.*

§ The process of systematically examining research evidence to assess its validity, results, and relevance before using it to inform a decision. This term is used for items 12 and 19 instead of "risk of bias" (which is more applicable to systematic reviews of interventions) to include and acknowledge the various sources of evidence that may be used in a scoping review (e.g., quantitative and/or qualitative research, expert opinion, and policy document).

*From:* Tricco AC, Lillie E, Zarin W, O'Brien KK, Colquhoun H, Levac D, et al. PRISMA Extension for Scoping Reviews (PRISMAScR): Checklist and Explanation. Ann Intern Med. 2018;169:467–473. [doi: 10.7326/M18-0850](http://annals.org/aim/fullarticle/2700389/prisma-extension-scoping-reviews-prisma-scr-checklist-explanation).
